# Factors associated with the stigma-discrimination complex towards healthcare workers among university students during the coronavirus pandemic in Mexico

**DOI:** 10.1101/2021.03.14.21253547

**Authors:** Carlos Arturo Cassiani-Miranda, Isabel Álvarez-Solorza, Adalberto Campo-Arias, Yinneth Andrea Arismendy-López, Andrés Felipe Tirado-Otálvaro, Lilia Patricia Bustamante-Montes, María Juana Gloria Toxqui-Tlachino

**Affiliations:** Associate professor, Programa de Medicina, Facultad de Ciencias de la Salud, Universidad de Santander (UDES), Bucaramanga, Colombia.; Associate professor, Facultad de Enfermería y Obstetricia de la Universidad Autónoma del Estado de México, Toluca, Mexico.; Associate professor, Programa de Medicina, Facultad de Ciencias de la Salud, Universidad del Magdalena, Santa Marta, Colombia.; Medical student, Programa de Medicina, Facultad de Ciencias de la Salud, Universidad de Santander (UDES). ( o); Titular professor, Facultad de Enfermería, Universidad Pontificia Bolivariana, Medellín, Colombia; Titular professor, Decanato de Ciencias de la Salud, Universidad Autónoma de Guadalajara, Guadalajara, México; Associate professor, Facultad de Enfermería y Obstetricia de la Universidad Autónoma del Estado de México, Toluca, Mexico

**Keywords:** Social Stigma, Risk Factors, Health Personnel, Students, COVID-19

## Abstract

The COVID-19-related stigma towards healthcare workers negatively influences their performance and job satisfaction, and well-being. The frequency of COVID-19-related stigma towards healthcare workers and its associated factors has not been sufficiently investigated. The objective was to determine the frequency and variables associated with COVID-19-related stigmatisation towards health workers in emerging-age university adults in Mexico. Analytical and cross-sectional study using an online questionnaire in 1,054 students between 18 and 29 years of age. Demographic variables, religiosity, fear of COVID-19 and stigma-discrimination related to COVID-19 towards healthcare workers were analysed. The latter was set as the dependent variable, while demographic variables, religiosity and high fear of COVID-19 were the independent variables. For the association between the variables, a binomial and logarithmic generalised linear model was designed to calculate the adjusted prevalence ratios. The proportion of high stigma-discrimination was 12.4%, and this was associated with a high fear of COVID-19 (APR 1.51, 95% CI 1.06 to 2.23). The main limitations were the cross-sectional nature, social desirability bias, non-probabilistic sampling. The results highlight the importance of establishing programmes to reduce COVID-19-related stigmatisation towards healthcare workers.

## Introduction

Infectious disease outbreaks are associated with negative psychological and social consequences comparable to their biological outcomes (Saeed et al., 2020). Among these consequences, one of the most impactful is stigma-discrimination, as it affects health-related outcomes and is a barrier to accessing health services (Asadi-Aliabadi et al., 2020). These negative social attitudes and behaviours affect various segments of society, such as patients, families, and healthcare providers (Asadi-Aliabadi et al., 2020).

During the rapid spread of SARS-CoV-2 infection, it has been observed that health systems have faced a critical overload on human resources (Sorokin et al., 2020), which are mainly due to psychological distress (Ramaci et al., 2020) associated with the so-called stigma-discrimination complex (SDC) (Campo-Arias & Herazo, 2015; Cassiani-Miranda et al., 2020).

Stigma is defined as a characteristic or trait attributed to someone or something and usually implies a negative connotation (Goffman, 2009). This trait represents a discrediting label that makes stigmatised people feel alienated from society, and labelling can turn into pigeonholing and stereotyping, leading to discrimination and loss of status (Link & Phelan, 2001).

In the context of the COVID-19 pandemic, people are stigmatised because of a perceived link to the disease, shaping the concept of COVID-19-related stigma, particularly affecting healthcare workers (HCWs) (Chopra & Arora, 2020). In infectious disease outbreaks, fear and misinformation are variables that explain stigmatisation (Schoch-Spana et al., 2010; Abuhammad et al., 2020). SDC interferes with the diagnosis and treatment process by disrupting communication, individual identity and sense of free will (Mawar et al., 2005). People who feel stigmatised tend to avoid certain behaviours perceived to increase stigma, such as refusing to be tested for COVID-19, because a positive result may be the label they consider stigmatising (Fischer et al., 2019).

Stigma is a barrier to effective infectious disease prevention and control mechanisms and can affect the stigmatised group and a wide range of people around them with a particular impact on HCWs (Perry & Donini-Lenhoff, 2010; Ramaci et al., 2020). For example, perceived stigma discourages infected persons and affects healthcare workers themselves (Almutairi et al., 2018) and can be an additional stressor (Ramaci et al., 2020).

Complementarily, negative attitudes may affect care-seeking and increase the collateral damage of COVID-19 (Cassiani-Miranda & Campo-Arias, 2020). More than 200 COVID-19-related attacks on HCWs have been documented (Bagcchi, 2020). In these incidents, HCWs experienced social isolation, public insults or harassment, refusal to use public transport and eviction from the housing. These violent behaviours in the pandemic context towards HCWs shape a specific stigmatisation process called Coronavirus Disease-Related Stigma-Discrimination-Complex-towards Healthcare Workers (CDRSDCHCW) (Campo-Arias et al., 2020; Stangl et al., 2019; Ransing et al., 2020).

These stigmatisation experiences negatively influence HCWs’ performance, job satisfaction, self-efficacy and general well-being (Monterrosa-Castro et al., 2020; Ramaci et al., 2020; Teksin et al., 2020). Besides, it is crucial to consider that SDC tends to persist over time, easier to consolidate than to eradicate (Dar et al., 2020). Therefore, CDRSDCHCW should be rigorously addressed by healthcare professionals, providers, and health authorities (Adiukwu et al., 2020). Having a valid and reliable instrument to measure CDRSDCHCW (Campo-Arias et al., 2020) could improve the identification of the phenomenon and associated variables and allow public health decision-makers to design informed and effective interventions. Despite the potential impact on the quality of life of HCWs, CDRSDCHCW and its associated factors have not been sufficiently investigated (Mostafa et al., 2020).

This study aimed to determine the frequency and explore variables associated with stigma discrimination complex related to coronavirus disease towards healthcare workers during the pandemic in a sample of emerging-age university adults in Mexico.

## Method

### Study design and ethical considerations

A research ethics committee approved this cross-sectional, analytical study of a Colombian state university (Act 004 of the extraordinary session of 13 May 2020). Participants gave informed consent in congruence with national and international standards (Colombian Ministry of Health, 1993; World Medical Association, 2018).

### Population and sample

An online questionnaire solicited the participation of students from a Mexican state university. A sample of at least 384 participants was expected since this sample size was calculated for low prevalences of the dependent variable of 10%, with a margin of error of 3%, or as high as 50%, with a margin of error of 5%; in both cases with a 95% confidence level (Hernández, 2006). This number of participants would explore some associations with acceptable confidence intervals (Katz, 2006). Students between 18 and 29 years of age (emerging age) were included (Arnett, 2000).

### Measures

The research questionnaire included demographic variables (age, gender, marital status, education level, occupation, marital status and income level), religiosity, fear of coronavirus disease and stigma-discrimination related to coronavirus disease towards HCWs.

### Stigma-discrimination complex related to coronavirus disease towards healthcare workers

It was quantified with the Stigma-Discrimination Scale Related to Coronavirus Disease towards Healthcare workers. This tool is a five-item instrument with a dichotomous response pattern (Campo-Arias et al., 2020), which resulted from a process of adaptation of the Tuberculosis Stigma Scale (Upegui-Arango & Orozco-Vargas, 2019). The instrument presents a unidimensional structure with acceptable internal consistency, Kuder-Richardson coefficient of 0.67 (Campo-Arias et al., 2020). Each affirmative response is assigned one point; overall scores can be between 0 and 5. Scores between 0 and 2 were classified as low stigma-discrimination related to coronavirus disease, and scores between 3 and 5 were classified as high stigma-discrimination according to the research group’s criteria. In the present study, the Kuder-Richardson test was 0.68.

### Religiosity

Religiosity was quantified with the Francis 4-item short scale of attitude towards Christianity (Francis-4). This instrument consists of four items exploring attitude towards God, Jesus and prayer. The items offer five response options from completely disagree to completely agree, scored from zero to four, with overall scores between 0 and 16. This instrument has shown excellent dimensionality and internal consistency in previous studies (Campo-Arias & Ceballos-Ospino, 2020). Global scores between 0 and 12 were rated as low religiosity and those between 13 and 16 as high religiosity, based on previous studies in Colombia (Campo-Arias et al., 2020). In the present study, the scale showed a Cronbach’s alpha of 0.97.

### Fear of coronavirus disease

It was quantified with the five-item version of the Fear of COVID-5 (Cassiani-Miranda et al., 2021) adapted and validated by the research team in the general Colombian population original 7-item Fear of COVID-19 Scale (Ahorsu et al., 2020). The Fear of COVID-5 has four response options that are scored from 0 to 3. The Spanish version of this instrument has shown adequate internal consistency, Cronbach’s alpha of 0.75 (Cassiani-Miranda et al., 2020). In the present study, the scale showed a Cronbach’s alpha of 0.78. The scale allows total scores between 0 and 15. Scores equal to or higher than four were categorised as high fear related to COVID-19, like a similar study in a Colombian population (Cassiani-Miranda et al., 2020).

### Procedure

An electronic questionnaire was sent via the institutional mail server. The invitation to participate in the study was sent to all persons linked to the university. The anonymous questionnaire took no more than ten minutes to complete. Data was collected between 3 July and 10 August 2020.

### Data analysis

Frequencies and percentages for qualitative variables and measures of central tendency and dispersion for quantitative variables [mean (M), standard deviation (SD), median (Me) and interquartile range (IQR)] were described according to the distribution of the data. For the analysis, CDRSDCHCW was considered the dependent variable, while demographic variables, religiosity, and high fear of COVID-19 were considered independent variables. For variables associated with CDRSDCHCW, prevalence ratios (PR) with their 95% confidence intervals (95% CI) were calculated. Subsequently, significant variables in the bivariate analysis, those that met the Hosmer-Lemeshow criteria (Hosmer et al., 1991) and those with biological plausibility with the outcome, were entered into a generalised linear model with a binomial distribution and log link function to calculate adjusted PRs (APRs) explaining the association between covariates and CDRSDCHCW (Bastos et al., 2015). The analysis was completed in Jamovi version 1.2.27.0 (Jamovi Project, 2020).

## Results

A total of 1,054 students participated (M = 20.7, SD = 2.2, Me = 20, IQR 19-22). Categorical findings and population characteristics are presented in table 1.

**Table 1.** Demographical characteristics of the sample.

| Variable | n | % |
| --- | --- | --- |
| Gender |  |  |
| Female | 852 | 80.8 |
| Male | 202 | 19.2 |
| Stable couple (marital status) |  |  |
| Yes (free union or married) | 58 | 5.5 |
| No (single, separated or widowed) | 996 | 94.5 |
| Have children |  |  |
| Yes | 60 | 5.7 |
| No | 994 | 94.3 |
| Bachelor's degree |  |  |
| Yes | 1046 | 99.2 |
| No | 8 | 0.8 |
| Employee |  |  |
| Yes | 156 | 14.8 |
| No | 898 | 85.2 |
| Healthcare worker |  |  |
| Yes | 171 | 16.2 |
| No | 883 | 83.8 |
| Low income (Mexican classification) |  |  |
| Yes | 448 | 42.5 |
| No | 606 | 57.5 |
| Low religiosity |  |  |
| Yes | 802 | 76.1 |
| No | 252 | 23.9 |
| Elevated fear COVID-19 |  |  |
| Yes | 254 | 24.0 |
| No | 801 | 76.0 |
| Stigma towards healthcare workers |  |  |
| Yes | 131 | 12.4 |
| No | 923 | 87.6 |

Scores for religiosity were observed between 0 and 16 (M = 8.53, SD = 5.45, Me = 9, IQR = 4-12); for fear of coronavirus disease they were between 0 and 15 (M = 2.35; SD = 2.51; Me = 2; and RIC = 0.25-3); for stigma-related discrimination between 0 and 5 (M = 0.92, SD = 1.26, Me = 0, IQR = 0-1). 12.4% (n=131) of participants scored for high stigma-discrimination. A statistically significant association was found between CDRSDCHCW and the high fear of COVID-19. See table 2.

**Table 2.** Associate factors with high stigma-discrimination in adult emergent students in México.

| Item | PR (95% CI) | APR (95%) |
| --- | --- | --- |
| Female gender | 1.32 (0.38-2.07) | 1.26 (0.82-2.05) |
| Stable couple | 0.97 (0.47-1.98) | 0.98 (0.38-2.14) |
| Have children | 0.94 (0.46-1.91) | 0.86 (0.33-1.85) |
| Bachelor's degree | 0.99 (0.16-6.16) | 0.96 (0.25-15.97) |
| Employee | 1.10 (0.71-1.70) | 1.09 (0.67-1.66) |
| Healthcare worker | 1.04 (0.68-1.60) | 0.97 (0.61-1.48) |
| Low income | 1.29 (0.94-1.78) | 1.33 (0.96-1.83) |
| Low religiosity | 0.83 (0.58-1.18) | 0.86 (0.61-1.25) |
| Elevated COVID fear | 1.55 (1.11-2.17) | **1.51 (1.06-2.23)** |

## Discussion

In the present study, 12.6% of Mexican emerging-age students reported high CDRSDCHCW and associated high fear of COVID-19. The high frequency of CDRSDCHCW observed is similar to that found in other studies that have reported high levels of COVID-19-related stigma towards healthcare workers (Abuhammad, 2020; Cassiani-Miranda et al., 2020; Sotgiu & Dobler, 2020). These findings could be explained by the fact that SARS-CoV-2 is more contagious than other stigmatised diseases (Schwartz et al., 2020). The perception of a global pandemic experience combined with economic crisis scenarios (Saeed et al., 2020) could also explain the high stigmatisation level.

The development of stigma/discrimination is related to cultural, educational, religious, personal, economic and environmental variables (Demirtaş-Madran, 2020). Thus, in the present study, a statistically significant association was observed between high CDRSDCHCW and a high level of fear of COVID-19. These results are compared with similar studies where fear of contagion is a variable that generates prejudice, stereotypes and discrimination (Abuhammad, 2020; Cassiani-Miranda et al., 2020). Indeed, HCWs around the world, in both low- and middle-income countries, face SDC, mediated mainly by fear of infection (Sotgiu & Dobler, 2020). However, the association between CDRSDCHCW and other variables may take on a different connotation according to methodological issues. For example, in a sample of 91 COVID-19 survivors in India, perceived stigma was significantly higher in men than women; enacted stigma and internalised stigma were associated with a high education and occupation level. (Dar et al., 2020). In this study, Dar et al. measured stigma using a stigma questionnaire adapted from the Ebola-related stigma questionnaire derived from the Berger HIV stigma scale (Jeyaseelan et al., 2013). However, unlike our study, the stigma assessment in this sample did not target stigmatisation experiences directed towards HCWs but was instead a general stigma measure, explaining the differences in associations with the present study.

Another predictor of the stigma that we included in this study was religiosity, although no statistical association with CDRSDCHCW was found. The influence of religiosity on COVID-19 management, especially in developing countries, may generate certain practices that are deleterious to established precautionary measures against the disease; furthermore, the belief that life and death are controlled by the “Almighty”, which is also configured as a religious stigma that may impact precautionary measures against COVID-19 (Hashmi et al., 2020). This observation suggests that the relationship between stigma and religiosity is more complex and may be mediated by other types of variables not explored in this study.

### Implications

The CDRSDCHCW could disrupt patient identification and surveillance and generate considerable negative impacts on pandemic control and management (Asadi-Aliabadi et al., 2020). Therefore, predicting the stigma-related consequences of COVID-19 is essential in planning prevention measures (Brooks et al., 2020; Liu et al., 2020). Consistent with previous data (Ramaci et al., 2020), this study’s findings suggest that the CDRSDCHCW study can provide insight into the stigmatisation process associated with emerging infectious diseases and the possible consequences of such stigmatisation.

Pandemic-related stigmatisation is a violation of human rights (Demirtaş-Madran, 2020) and undermines basic levels of dignity, provokes new types of vulnerabilities, and reinforces pre-existing inequalities. (Ugidos et al., 2020; Ramaci et al., 2020). Therefore, knowing the frequency and correlates of CDRSDCHCW provides valuable information for designing interventions to reduce social stigma towards HCWs (Chopra & Arora, 2020; Demirtaş-Madran, 2020). In this perspective, an inclusive and rights-based policy approach to address COVID-19 should not only focus on eliminating the disease or interrupting transmission but also address stigma and intersecting vulnerabilities, as well as the conditions that promote or perpetuate stigma (Bologna et al., 2021; Qin X & Song, 2021). Consequently, to be equitable and practical, the global response to COVID-19 requires that stigma becomes a higher priority on the public health agenda (Roelen et al., 2020).

### Limitations

The cross-sectional nature of this study limits the strength of the associations assessed, and it is not possible to make causal relationships between variables. On the other hand, social desirability bias may have limited participants’ responses for fear of being stigmatised, although to reduce this, we tried to preserve maximum confidentiality. The results are not generalisable to the Mexican university population, as the convenience sample was restricted to a single university. The study’s quantitative nature does not allow us to explore other manifestations of the stigmatisation process related to COVID-19 so that qualitative studies would enrich the state of knowledge in this area.

## Conclusions

The COVID-19-related stigmatisation towards health workers among emerging-age adult university students in Mexico is considerable and is associated with a high level of fear of COVID-19. These results highlight the importance of establishing intervention programmes that address COVID-19-related stigma towards healthcare workers and their potential consequences.

## Data Availability

The data that support the findings of this study are available from the corresponding author upon reasonable request.

